# Aberrant effective connectivity is associated with positive symptoms in first-episode schizophrenia

**DOI:** 10.1101/19012773

**Authors:** Martin J. Dietz, Yuan Zhou, Lotte Veddum, Christopher D. Frith, Vibeke F. Bliksted

## Abstract

Schizophrenia is a tenacious psychiatric disorder thought to result from synaptic dysfunction. While symptomatology is traditionally divided into positive and negative symptoms, abnormal social cognition is now recognized a key component of schizophrenia. Nonetheless, we are still lacking a mechanistic understanding of how aberrant synaptic connectivity is expressed in schizophrenia during social perception and how it relates to positive and negative symptomatology. We used fMRI and dynamic causal modelling (DCM) to test for abnormalities in synaptic efficacy in twenty-four patients with first-episode schizophrenia (FES) compared to twenty-five matched controls performing the Human Connectome Project (HCP) social cognition paradigm. Patients had not received regular therapeutic antipsychotics, but were not completely drug naïve. Our data reveal an increase in excitatory feedforward connectivity from motion-sensitive V5 to posterior superior temporal sulcus (pSTS) in patients compared to matched controls. At the same time, were less accurate than controls in judging social stimuli from non-social stimuli. Crucially, patients with a higher degree of positive symptoms had more disinhibition within pSTS, a region computationally involved in Theory of Mind. We interpret these within a predictive coding framework where increased feedforward connectivity may encode aberrant prediction errors from V5 to hierarchically higher pSTS and local disinhibition within pSTS may reflect aberrant encoding of the precision of cortical representations about social stimuli.

## Introduction

Schizophrenia is a tenacious psychiatric disorder that affects about 1% of the population worldwide (Javitt, 2010). The first symptoms tend to manifest in the mid-twenties in the form of a psychotic episode. While symptomatology is traditionally divided into positive and negative symptoms, abnormal social cognition is recognized a key component of schizophrenia (Green et al., 2015). Recent research has shown that patients with high levels of both positive and negative symptoms, or negative symptoms alone, have more profound deficits in theory of mind compared to patients with only positive symptoms (Kästner et al., 2015; Bliksted et al., 2016). While negative symptoms have been associated with hypo-mentalizing similar to autism (Bliksted et al., 2016), positive symptoms have been associated with hyper-mentalizing, in particular paranoia (Ciaramidaro et al., 2015). Importantly, it has been suggested that patients with schizophrenia could be switching between hyper- and hypo-mentalizing (Bliksted et al., 2016; 2018). Theory of mind refers to an agent’s ability to generate an internal model of another agent’s beliefs about the world. From a computational perspective, this internal model is necessary to explain and predict other agents’ behavior in terms of their intentions, goals and desires (Friston and Frith, 2015). fMRI studies in healthy individuals have consistently associated theory of mind with the posterior superior temporal sulcus (pSTS) and the medial prefrontal cortex (Blakemore, 2008; Green et al., 2015). One of the most validated (Blakemore, 2008; Pinkham and Harvey, 2013) and widely used theory-of-mind tasks is the Animated Triangles Task (ATT) (Abell et al., 2000). This paradigm has also been used to identify abnormal BOLD activation in patients with schizophrenia compared to healthy controls (Pedersen et al., 2012; Das et al., 2012a; 2012b; Koelkebeck et al., 2013; Bliksted et al., 2018). However, results have been inconsistent. While Das *et al*. found reduced activation of the right superior temporal gyrus (STG), the temporo-parietal junction (TPJ) and bilateral inferior frontal gyri (IFG) (Das et al., 2012a; 2012b), Martin *et al*. showed increased activation of bilateral IFG, left STG and left caudate nucleus (Martin et al., 2016). Finally, Bliksted *et al*. found that patients hyper-mentalized during non-social stimuli, accompanied by increased activation of the anterior medial prefrontal cortex (Bliksted et al., 2018).

Etiologically, schizophrenia is a neurodevelopmental disorder that is thought to emerge with synaptic dysfunction (Adams et al., 2013). In addition synaptic abnormalities, there is evidence of widespread white-matter abnormalities that could impair axonal conduction between brain areas (Shepherd et al., 2012; Yao et al., 2013; Kelly et al., 2018). Several studies have used non-invasive electrophysiology (EEG and MEG) to identify abnormal post-synaptic responses to both auditory (Umbricht and Krljes, 2005; Ranlund et al., 2015; Avissar et al., 2018), visual (Tsuchimoto et al., 2011; Sun et al., 2013; Tan et al., 2013; Grent-’t-Jong et al., 2016) and tactile (Reite et al., 2003; Huang et al., 2009) stimuli in patients with schizophrenia compared to healthy controls. Nonetheless, we are still lacking a more mechanistic understanding of how the effective brain connectivity that supports social perception is expressed in patients with first-episode schizophrenia. More specifically, we want to identify how aberrant effective connectivity during social perception is differentially related to positive and negative symptomatology. To this end, we used functional magnetic resonance imaging (fMRI) during the HCP social cognition paradigm (Barch et al., 2013) to test for abnormalities in effective connectivity in twenty-four patients with first-episode schizophrenia (FES) compared to twenty-five matched controls.

## Methods and Materials

### Patients

We initially recruited 31 patients from the OPUS Clinic, a first-episode schizophrenia clinic at Aarhus University Hospital, Denmark. While each patient met the ICD-10 criteria for schizophrenia, they had no history of neurological disorder or severe head trauma according to ICD-10, nor did they have an ICD-10 diagnosis of drug- or alcohol dependency. Patients were excluded if they had an estimated premorbid IQ < 70 based on their history or if they were not able to understand spoken Danish sufficiently well to comprehend the testing procedures. Seven patients were excluded from the fMRI experiment due to dental braces (N = 3), pregnancy (N = 1) and several no-shows (N = 3). Finally, 24 first-episode patients were included in the fMRI experiment. Given that our patients were newly diagnosed, most of them did not receive regular doses of antipsychotic medication at a therapeutic level that could be converted to standard chlorpromazine equivalents (Woods, 2005). Most of had only received depot injections or very low doses of antipsychotics used as an *ad hoc* sedative. Table 1 summarizes the cohort’s medication histories.

**Table 1.**
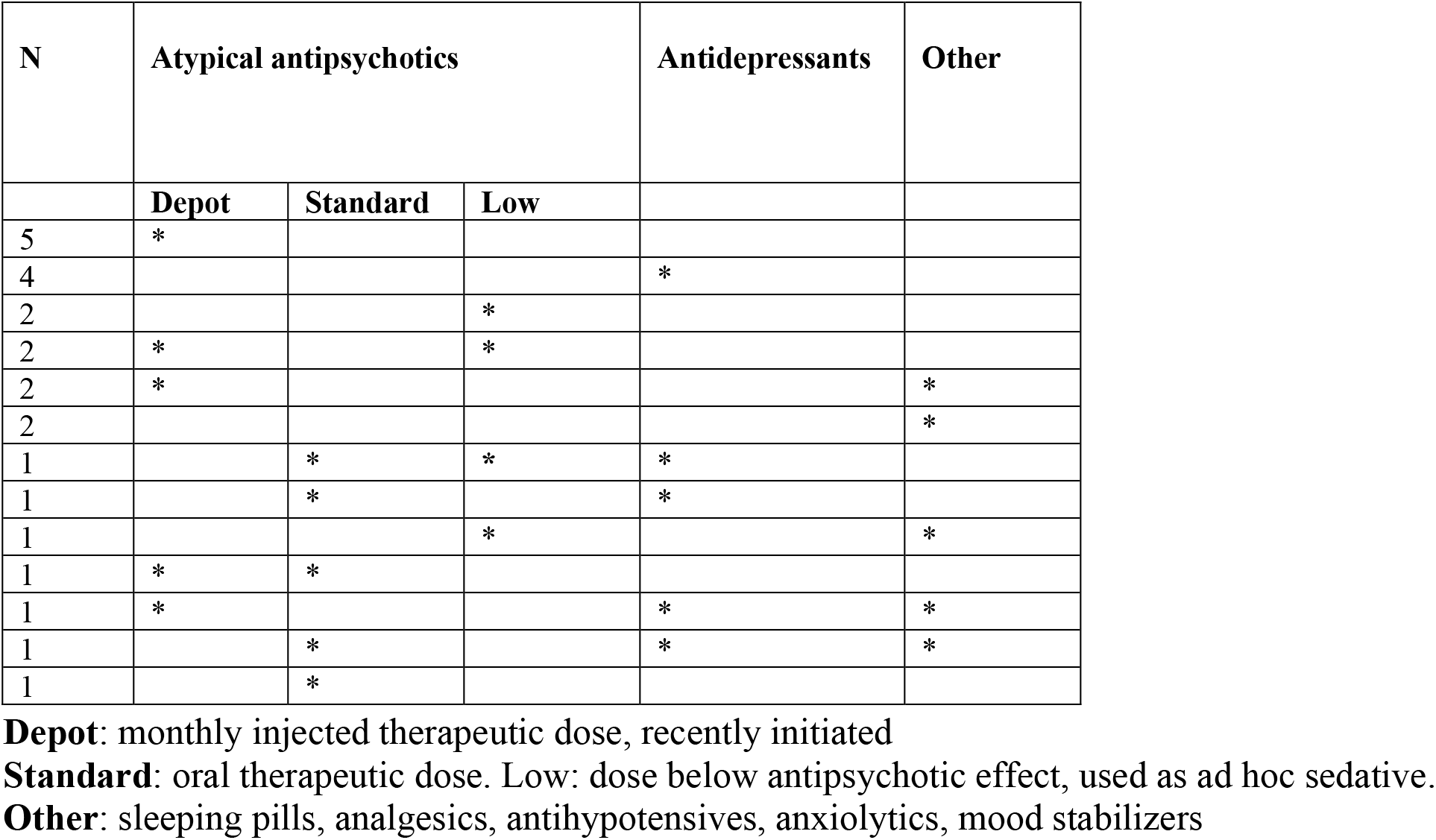
Medication history of first-episode schizophrenia patients

| N | Atypical antipsychotics |  |  | Antidepressants | Other |
| --- | --- | --- | --- | --- | --- |
|  | Depot | Standard | Low |  |  |
| 5 | * |  |  |  |  |
| 4 |  |  |  | * |  |
| 2 |  |  | * |  |  |
| 2 | * |  | * |  |  |
| 2 | * |  |  |  | * |
| 2 |  |  |  |  | * |
| 1 |  | * | * | * |  |
| 1 |  | * |  | * |  |
| 1 |  |  | * |  | * |
| 1 | * | * |  |  |  |
| 1 | * |  |  | * | * |
| 1 |  | * |  | * | * |
| 1 |  | * |  |  |  |

**Depot:** monthly injected therapeutic dose, recently initiated
**Standard:** oral therapeutic dose. Low: dose below antipsychotic effect, used as ad hoc sedative.
**Other:** sleeping pills, analgesics, antihypotensives, anxiolytics, mood stabilizers

### Healthy controls

We initially recruited 29 healthy controls. Exclusion criteria were the same as for the patients, except that controls were excluded if they or a first-degree relative had an ICD-10 diagnosis, or if a diagnosis was confirmed during the PSE interview. Four controls were excluded from the fMRI experiment on the day of scanning: two had dental braces and two left the study prematurely. FES patients and healthy controls were intended matched on age, gender, educational level (last commenced education), and parental socioeconomic status (SES). However, we did not succeed completely with this strategy, so the two groups ended up being matched on a group level. Finally, 25 healthy controls were included in the fMRI study.

### Psychopathology and social functioning

FES patients were interviewed by a psychiatrist with the Present State Examination (PSE) interview (ICD-10, WHO) regarding schizophrenia and drug dependency (World Health Organization, 1994). The healthy controls were interviewed with the entire PSE interview. All FES patients and healthy controls were rated with the Scale for the Assessment of Negative symptoms (SANS) and the Scale for the Assessment of Positive Symptoms (SAPS) (Andreasen, 1984a; 1984b). Level of psychosocial functioning was measured using the Global Assessment of Functioning (GAF-F) (Startup et al., 2002) and the Personal and Social Performance Scale (PSP) (Nasrallah et al., 2008).

### Behavioural tests

We estimated intelligence using two subtests from the Wechsler Adult Intelligence Scale (WAIS-III) (D, 1997). The two subtests were chosen based on their high correlation with the total WAIS-III IQ-score: Block Design and Vocabulary. Theory of mind ability was measured using the Animated Triangles Task (ATT) (Abell et al., 2000). In the “random” condition, the triangles move randomly about. In the theory of mind condition, the animated triangles move in a coordinated fashion that resembles a social interaction, a scenario that normally developing individuals consistently explain using theory of mind (Abell et al., 2000). There are four clips of each type of animation with 38-41 seconds duration. After each animation, the participants were asked to describe what they thought was happening and their answers were scored regarding degree of mental state attribution (range 0-5) and appropriateness of their description (range 0-3) as outlined by Castelli et al. (37). Each answer was evaluated by two clinical psychologists (LV and VB). Intraclass correlations in a two-way random-effects model showed absolute agreement of intentionality scores in the “random” condition (ICC = 0.97, 95% CI (0.93; 0.99), *P*<0.0001) and the “theory of mind” condition (ICC = 0.96 95% CI (0.89; 0.98), *P*<0.001) and absolute agreement of appropriateness scores in the “random” condition (ICC = 0.97, 95% CI (0.92; 0.99), *P*<0.001) and the “theory of mind” condition (ICC = 0.96, 95% CI (0.91; 0.98), *P*<0.001) among the two ratings.

### fMRI paradigm

We used the social cognition paradigm from the Human Connectome Project (HCP) (Barch et al., 2013) with permission from the WU-Minn HCP consortium (http://www.humanconnectome.org/). This paradigm consists of animated sequences of 20 seconds duration showing geometric shapes (triangles, squares and circles) that move about in either a coordinated fashion that resembles a social interaction among individuals (social motion) or in a random fashion (non-social motion). Participants were presented with ten sequences of social scenarios and ten sequences of random scenarios in randomized order. After each stimulus sequence, participants were asked whether they had perceived a social interaction, answering ‘yes’ with their right index finger or ‘no’ with their right middle finger (see Fig. 1). The paradigm was presented with E-prime 2.0 (Psychology Software Tools, Inc.) and projected onto a screen at the back of the MRI bore that participants viewed through a mirror mounted on top of the radio frequency coil. Each block of stimulus and response was followed by 15 seconds of visual fixation. The total duration of the paradigm was 12.54 minutes.

**Figure 1.**
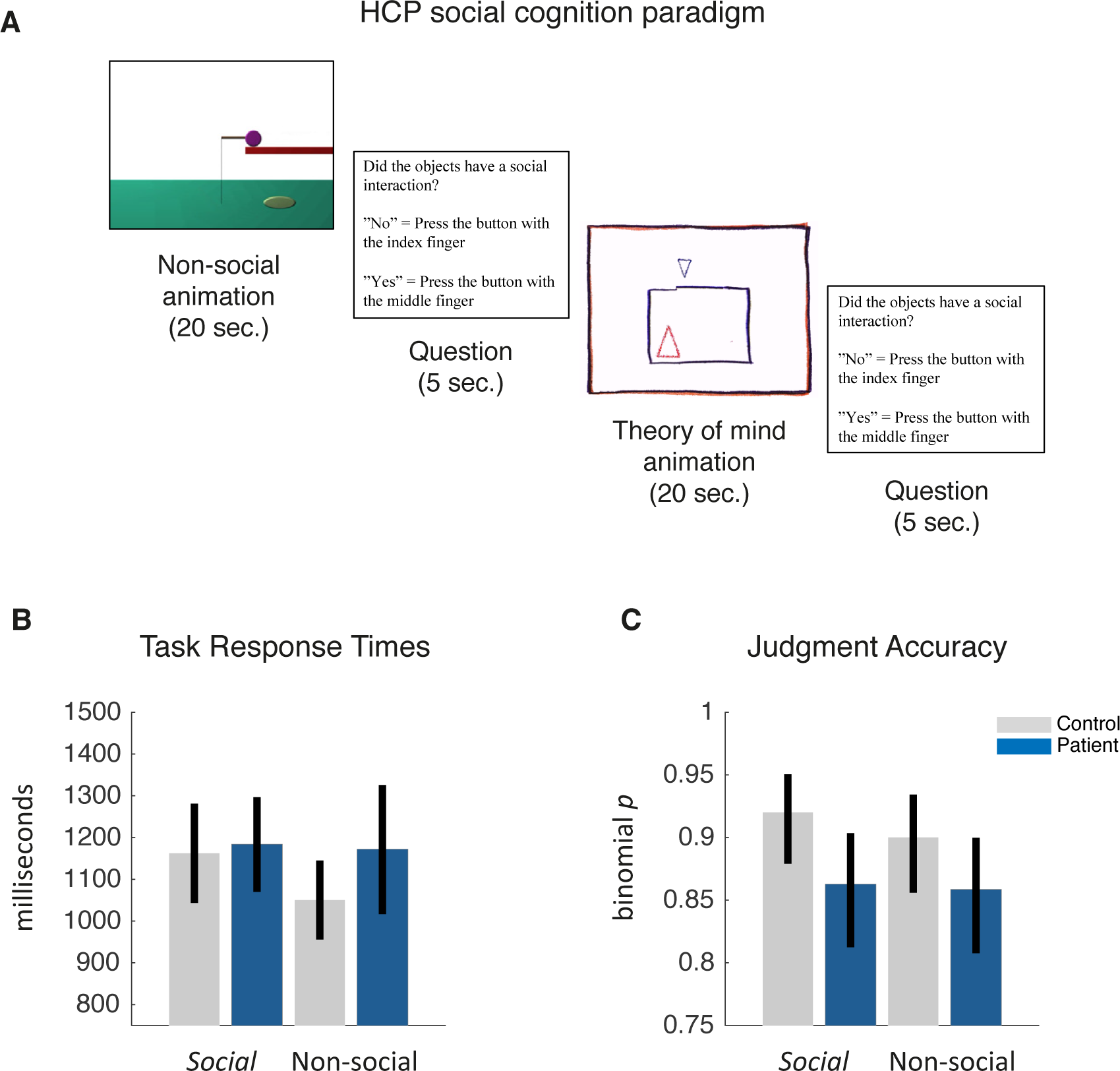
Judgment of social versus non-social scenarios **(A)** Social cognition paradigm **(B)** Response times with 95% confidence intervals **(C)** Judgment accuracy with 95% confidence intervals: patients with schizophrenia were less accurate in detecting social scenarios than healthy controls, with no consistent difference between groups when judging non-social scenarios.

### Ethics statement

All participants received written and verbal information about the project and a written informed consent was obtained before inclusion. The study was approved by the Central Denmark Region Committee on Health Research Ethics (Ref: 1-16-02-87-15) and the Danish Data Protection Agency. The project complied with the Helsinki-II-declaration.

### fMRI data acquisition

T2*-weighted echo planar images (EPI) were acquired on a 3T Siemens Magnetom Trio using a 32-channel radio frequency head coil. Each volume consisted of 40 slices with 3 mm thickness acquired in descending order with repetition time (TR) = 2 sec, echo time (TE) = 27 ms, flip angle = 90°, field of view (FOV) = 192 × 192 mm and in-plane resolution = 64 × 64.

### fMRI analysis

fMRI data were analyzed using Statistical Parametric Mapping (SPM12, revision 6906). Images were resampled to 2 mm^3^ voxels, realigned and spatially normalized to MNI space. Time-series were high-pass filtered at 1/128 s using a discrete cosine set and temporal correlations were modelled using an AR(1) model. Social and non-social motion conditions were modelled as boxcar regressors convolved with a canonical HRF and fitted to the BOLD time-series using a general linear model (Worsley and Friston, 1995). Visual fixation periods were not modelled and hence constituted an implicit baseline. Finally, the realignment parameters were modelled to adjust for the effects of head movement. We created contrast images for each patient and control testing for visual motion in general (both social and non-social versus fixation) and the difference in activation between social and non-social motion. Contrast images were smoothed with a 6-mm FWHM Gaussian kernel and used as summary statistics in a random-effects analysis using one-sample *t*-tests within patients and controls, separately. To identify regions active both during the perception of visual motion in general and during social motion in particular, we used a conjunction analysis to test for a conjunction of *t*-tests (global null). This corresponds to masking one significant contrast with another to identify an overlap of significant activations (Friston et al., 2005). We thus tested for visual motion in general (conjunction) and for social compared to non-social stimuli to identify effects in each group separately, differences between groups and, finally, commonalities across groups. All statistical tests were thresholded at *p* < 0.05, family-wise error (FWE) whole-brain corrected for multiple comparisons using random field theory (Worsley et al., 1996).

### Dynamic causal modelling of effective connectivity

We used a two-state dynamic causal model (DCM) for fMRI (DCM12, revision 6755) to estimate the effective connectivity within and between brain areas, given observed haemodynamic measurements (Friston et al., 2003). While one-state DCM for fMRI is used to model extrinsic connections only, two-state DCM models both extrinsic connections between regions as excitatory (glutamatergic) forward and backward connections and intrinsic connectivity within each region in terms of one inhibitory (GABAergic) population and one excitatory (glutamatergic) population of neurons. This allows us to model the intrinsic connectivity within each cortical area as an increase or decrease in cortical inhibition (Marreiros et al., 2008). We summarised the BOLD signal in each participant using the first eigenvariate (principal component) of voxels within a sphere of 8 mm radius centred on each participant’s local maximum. This subject-specific local maximum was identified within a sphere of 20 mm radius centred on the peak of the group effect. The network derived empirically from the group-level fMRI result comprised motion-sensitive area V5 and pSTS in the right hemisphere (see Table 3.). Hemodynamic responses to all visual motion (social and non-social scenarios) were modelled as a driving input to area V5. Using parametric modulation of the regressor encoding all visual motion, responses to social compared to non-social biological motion were modelled as a modulatory input to the network under four alternative hypotheses. The first hypothesis was formulated as a full DCM, where both extrinsic (excitatory) connections between V5 and pSTS and intrinsic (inhibitory) connections within V5 and pSTS encode the differences between experimental conditions. The second hypothesis was formulated as a reduced model where only extrinsic (excitatory) connections between V5 and pSTS encode the differences between experimental conditions. The third hypothesis was formulated as another reduced model where only the forward (excitatory) connection from V5 and pSTS encodes the differences between experimental conditions. Finally, the fourth hypothesis encodes the belief that no connections encode any differences between conditions. (see Figure 3 for a schematic of alternative hypotheses). The posterior probability of the free parameters and the Bayesian model evidence of each model was then estimated using Bayesian model reduction (Friston et al., 2015).

**Figure 2.**
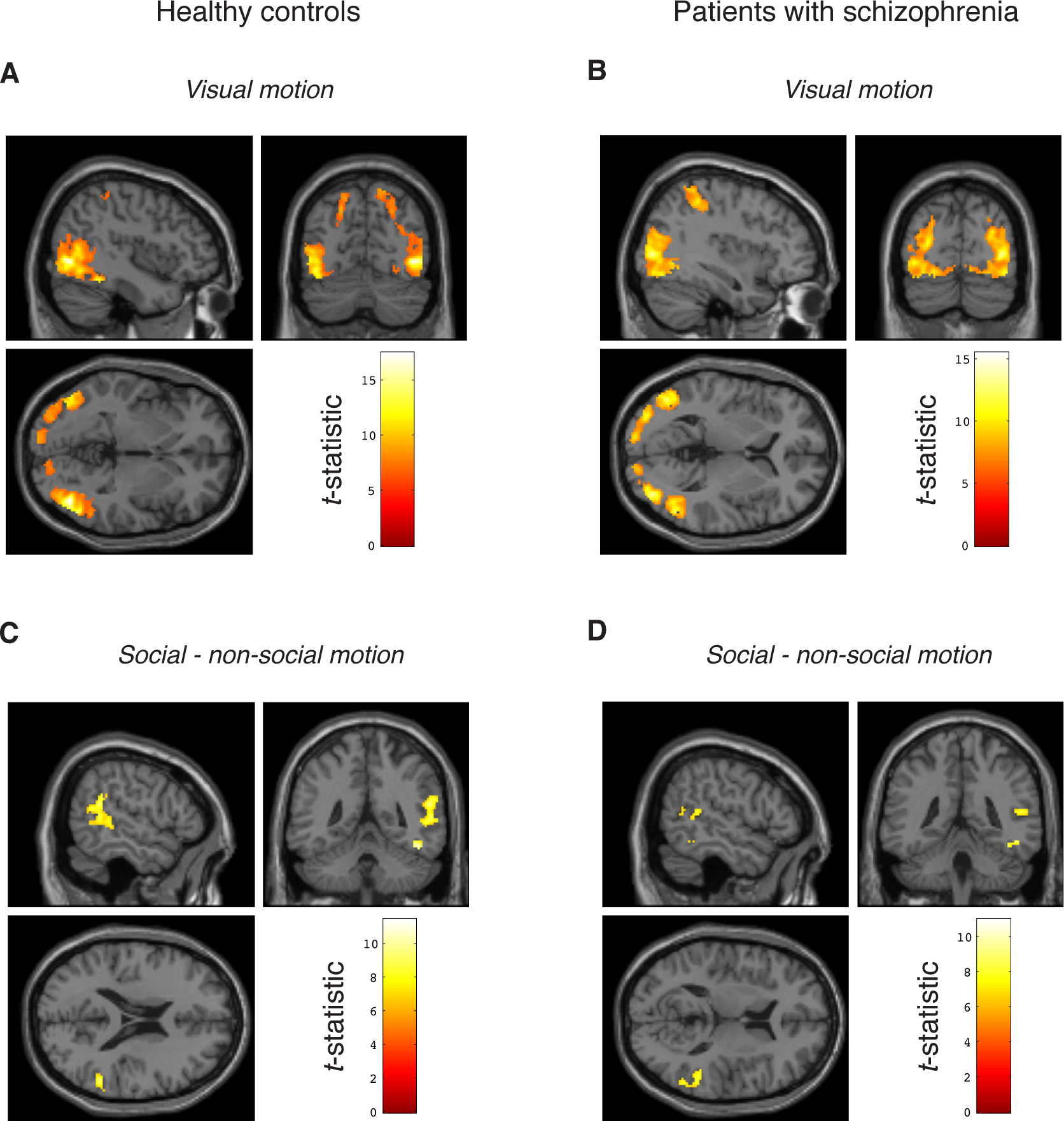
fMRI brain mapping in healthy controls and FES patients **(A)** Visual motion in healthy controls **(B)** Visual motion in patients with schizophrenia **(C)** Social minus non-social motion in healthy controls **(D)** Social – non-social motion in patients with schizophrenia. Statistical *t*-maps are thresholded at *p* < 0.05, FWE-corrected for multiple comparisons and rendered on a single-subject structural MRI in MNI space. See main text for MNI coordinates.

**Figure 3.**
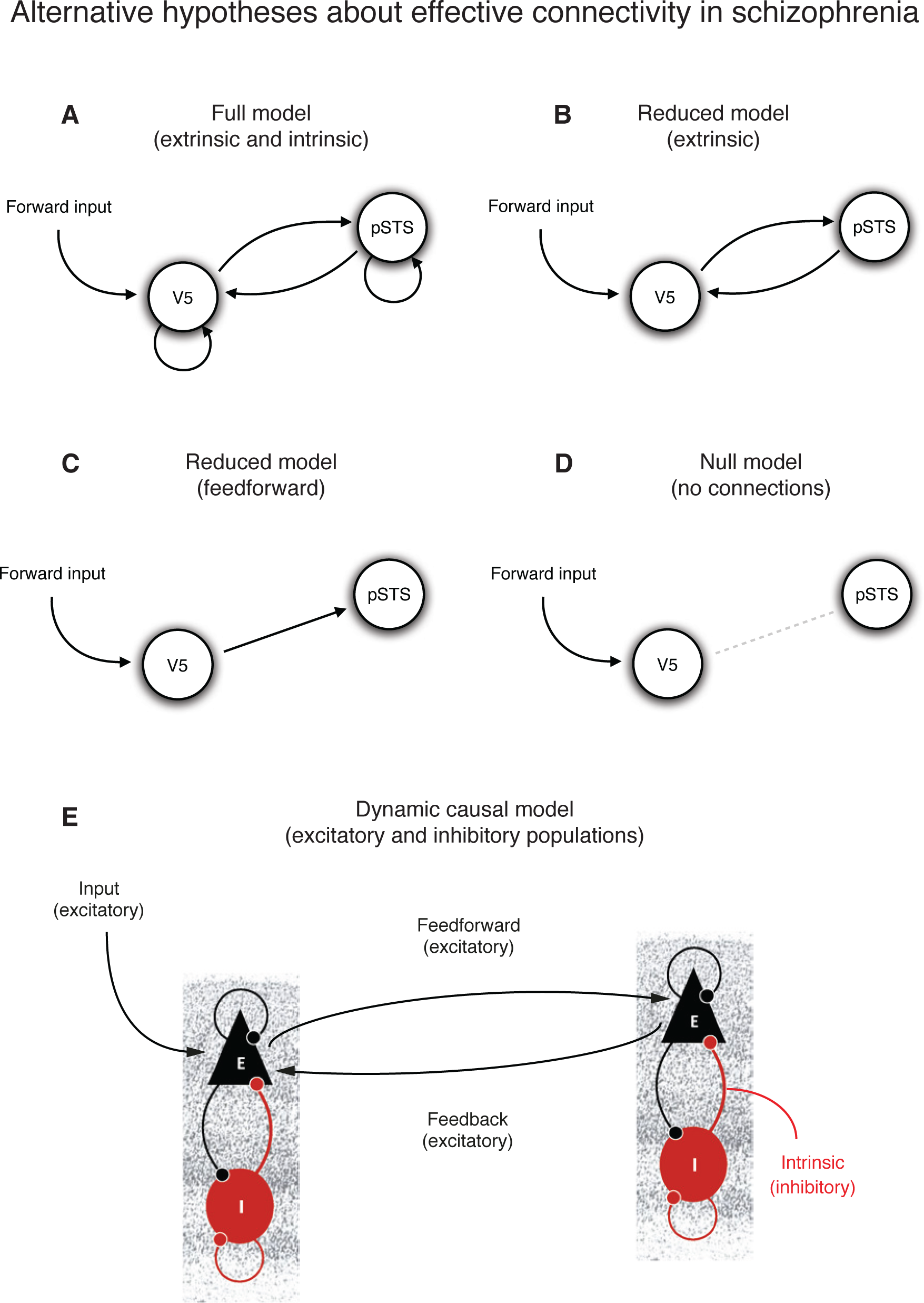
Alternative hypotheses about changes in effective connectivity during theory of mind in patients with schizophrenia **(A)** Full model with free parameters on extrinsic and intrinsic connections **(B)** Reduced model with free parameters on extrinsic connections **(C)** Reduced model with a free parameter the on feedforward connection only **(D)** Null model with all connections switched off (**E**) Schematic of the two-state DCM used to model each cortical region in terms of one excitatory (E) and one inhibitory (I) population of neurons that are coupled within each region via excitatory (black) and inhibitory (red) connections. The extrinsic connectivity between regions is modelled via excitatory forward and backward connections.

**Figure 4.**
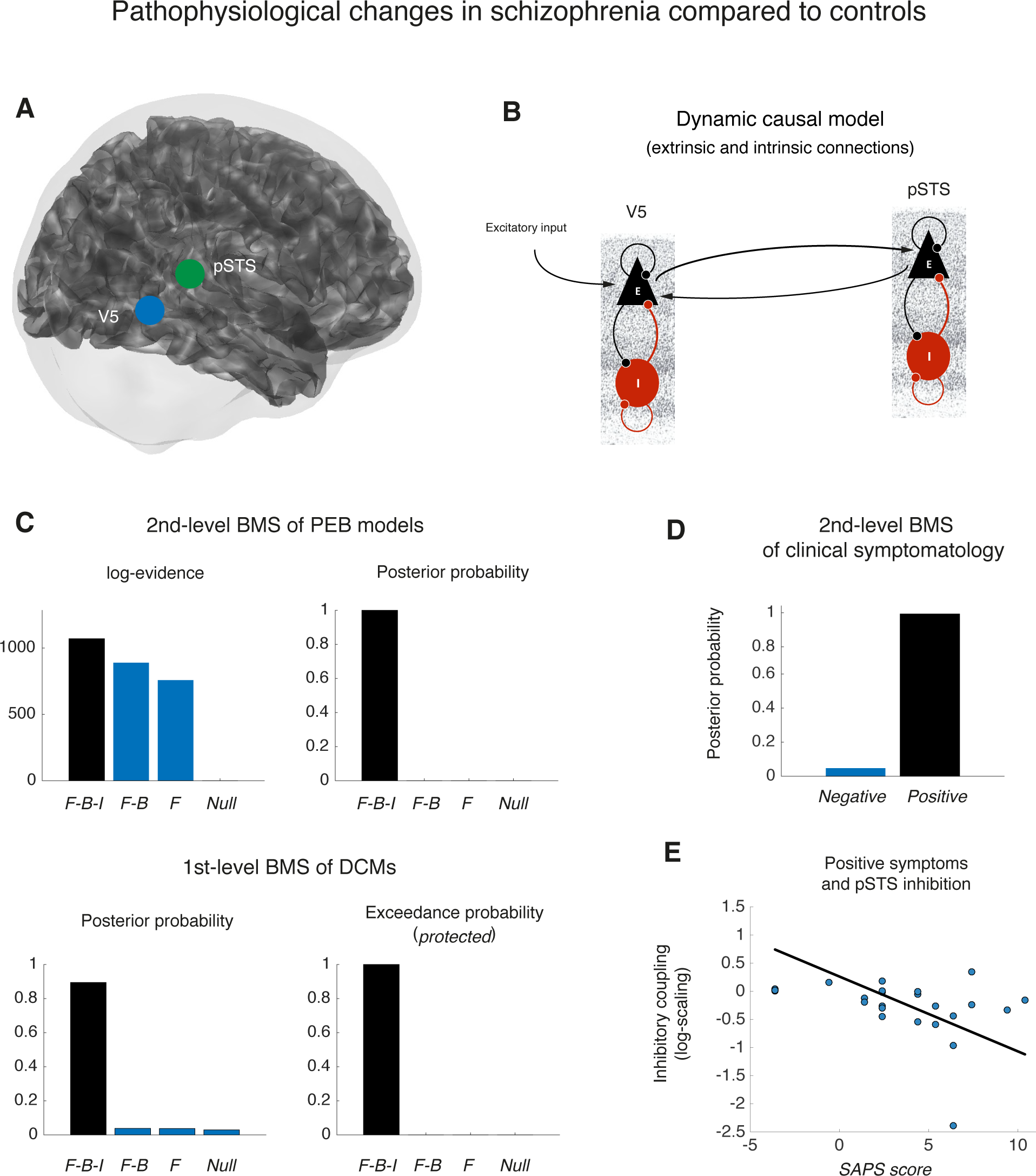
Pathophysiology in first-episode schizophrenia compared to healthy controls **(A)** Anatomical regions-of-interest used in dynamic causal modelling derived from MNI coordinates in Table 3. **(B)** Schematic of the full DCM that best explains the differences between patients and healthy controls **(C)** Bayesian model selection of first-level log-evidences of each participants’ DCMs and second-level log-evidences of PEB models at the group level. There is agreement between BMS at both levels that the full model with forward, backward and intrinsic (*F-B-I*) connections has high posterior probability compared to the reduced models with only forward and backward connections (*F-B*), feedforward coupling (*F*) or null model **(D)** Bayesian model selection of PEB models of clinical symptomatology, showing that positive symptoms are a better explanation of individual differences in effective connectivity than negative symptoms in this cohort **(E)** Patients with stronger positive symptoms had more disinhibition within pSTS.

### Parametric Empirical Bayesian analysis of group effects

We then used parametric empirical Bayes (PEB) to identify increases or decreases in extrinsic (excitatory) connections between V5 and pSTS and intrinsic (inhibitory) connections within each region at the group level. PEB is a hierarchical Bayesian model in which empirical priors on the connection strengths at the single-subject level are estimated empirically using a Bayesian general linear model at the group level. PEB allows us to identify differences in connection strengths between patients and healthy controls at the group level directly. In addition, using Bayesian model comparison of PEB models, we compared two alternative hypotheses about the association between clinical symptomatology and effective connectivity using Bayesian linear regression. In this way, we are able to disambiguate between positive and negative symptoms as the best explanation of patient variability in synaptic efficacy. Normally, one would include medication dose for antipsychotics as a nuisance regressor in the regression model. However, given that all patients were newly diagnosed and did not yet receive standard antipsychotic treatment at a therapeutic level, conversion of their heterogeneous medication to standard chlorpromazine equivalents was not feasible (Woods, 2005). Hence, we were not able to reliably adjust the regression models of positive and negative symptoms for standard doses of antipsychotics.

## Results

### Demographics, psychopathology, IQ and social cognition

Given that patients and controls were matched with regard to age, gender, educational level (last commenced education), and parental socioeconomic status (SES), we did not observe differences between groups in estimated IQ and, to our surprise, no differences in theory of mind (see Table 2).

**Table 2.** Demographics, psychopathology, IQ and social cognition

|  | **Schizophrenia**<br>**(N=24)** | **Healthy controls**<br>**(N=25)** | **Statistic**<br>**(degrees of**<br>**freedom)** | **P-value** |
| --- | --- | --- | --- | --- |
| **Age - mean (95% CI)** | 25.21 (23.35-27.07) | 24.6 (22.78-26.42) | $z = -0.62$ | 0.53 |
| **Females - N (%)** | 7 (29) | 11 (46) | $\chi^2(1) = 1.16$ | 0.28 |
| **Years of education**<br>**- mean (95%CI)** | 15.9 (14.91-16.89) | 14.60 (13.59-15.62) | $z = 1.61$ | 0.11 |
| **Current occupation - N (%)** | | | $\chi^2(5) = 18.06$ | 0.003<br>(0.001) |
| **Unemployed** | 6 (25) | 1 (4) |  |  |
| **Work** | 7 (29) | 5 (20) |  |  |
| **Student** | 5 (21) | 19 (76) |  |  |
| **Sick leave** | 3 (13) | 0 |  |  |
| **Pension** | 1 (4) | 0 |  |  |
| **Other** | 2 (8) | 0 |  |  |
| **SANS** | 8.17 (7.02-9.31) | 1.32 (0.20-2.44) | $z = -5.55$ | <0.0001 |
| **SAPS** | 7.08 (5.97-8.20) | 0.28 (-0.81-1.37) | $z = -5.54$ | <0.0001 |
| **PSP** | 55.03 (48.93-61.23) | 86.32 (83.20-89.44) | $t(34) = 9.37$ | <0.001 |
| **GAF-F** | 56.39 (51.78-61.00) | 86.56 (82.14-90.98) | $z = 5.56$ | <0.001 |
| **WAIS-III (estimated IQ)** | 92.96 (84.49-101.43) | 97.4 (86.19-108.61) | $t(47) = 0.65$ | 0.52 |
| **Intentionality ToM (ATT)** | 14 (12.67-15.33) | 15.24 (13.94-16.54) | $z = 1.31$ | 0.19 |
| **Intentionality Random (ATT)** | 0.75 (0.28-1.22) | 1.67 (0.64-2.70) | $z = -1.58$ | 0.11 |
| **Accuracy ToM (ATT)** | 8.04 (7.26-8.82) | 8.88 (8.12-9.64) | $z = 1.47$ | 0.14 |
| **Accuracy Random (ATT)** | 0.75 (0.26-1.24) | 2.27 (0.83-3.71) | $z = -1.96$ | 0.05 |

**SANS:** Scale for Assessment of Negative Symptoms **SAPS:** Scale for Assessment of Positive Symptoms **PSP:** Personal and Social Performance scale **GAF-F:** Global Assessment of Functioning – level of social functioning **WAIS-III:** Wechsler Adult Intelligence Scale-III (Block Design, Vocabulary) **ATT:** Animated Triangles

### Behavioural results of HCP fMRI paradigm

In the fMRI experiment, we saw that controls were slower in responding during social compared to non-social stimuli (*t*(24) = 2.26, *p* = 0.01, two-tailed *t*-test). In contrast, patients with schizophrenia showed no difference when judging between social and non-social biological motion (*t*(23) = −0.35, *p* = 0.7, two-tailed *t*-test). When comparing patients and controls, we found no evidence of a difference in response times for social (*t*(47) = −0.25, *p* = 0.8, two-tailed *t*-test), nor non-social motion (*t*(47) = −1.6, *p* = 0.1, two-tailed *t*-test). We then tested for a difference in task accuracy using McNemar’s *Chi*^*2*^-test. This tests for a difference in proportions of accurately judged scenarios within each group, separately. Neither control subjects (*Chi*^*2*^(1) = 1.08, *p* = 0.3), nor patients with schizophrenia (*Chi*^*2*^(1) = 0.03, *p* = 0.9) showed evidence of a difference in judgment accuracy between social and non-social stimuli. We then tested for a difference in judgment accuracy between patients and controls using Pearson’s *Chi*^*2*^-test. This revealed that controls were more accurate in judging social motion than patients with schizophrenia (*Chi*^*2*^(1) = 4.19, *p* = 0.04). In contrast, there was no evidence of a difference in accuracy between patients and controls when judging non-social motion (*Chi*^*2*^(1) = 2, *p* = 0.2). (see Figure 1.)

### fMRI results in healthy controls

When healthy controls perceived motion in general (social and non-social) they had increased activation in motion-sensitive area V5 in the right hemisphere with peak at MNI coordinate [44 −70 −2], *T*(24) = 17.41, *P*_*FWE*_ < 0.0001, as well as V5 in the left hemisphere with peak at MNI coordinate [−44 −72 −10], *T*(24) = 13.42, *P*_*FWE*_ < 0.0001. We also observed increased activation in the left superior parietal lobule (SPL) with peak at MNI coordinate [−30 −50 56], *T*(24) = 7.69, *P*_*FWE*_ < 0.0001. In contrast, when healthy controls perceived social motion compared to non-social motion, they had increased activation in posterior inferior temporal gyrus with peak at MNI coordinate [48 −50 −18], *T*(24) = 11.40, *P*_*FWE*_ < 0.0001 and posterior superior temporal sulcus (pSTS) with peak at MNI coordinate [56 −46 20], *T*(24) = 9.40, *P*_*FWE*_ < 0.0001.

### fMRI results in patients with first-episode schizophrenia

When patients with first-episode schizophrenia perceived motion in general (social and non-social) they had increased activation in motion-sensitive area V5 in the right hemisphere with peak at MNI coordinate [38 −80 4], *T*(23) = 13.40, *P*_*FWE*_ < 0.0001, as well as V5 in the left hemisphere with peak at MNI coordinate [−48 −72 0], *T*(23) = 15.45, *P*_*FWE*_ < 0.0001. We also observed increased activation in the right superior parietal lobule (SPL) with peak at MNI coordinate [10 −58 60], *T*(23) = 7.24, *P*_*FWE*_ < 0.0001. In contrast, when patients perceived social biological motion compared to non-social motion, they had increased activation in posterior inferior temporal gyrus with peak at MNI coordinate [46 −52 −14], *T*(23) = 9.30, *P*_*FWE*_ < 0.0001 and posterior superior temporal sulcus (pSTS) with peak at MNI coordinate [54 −42 12], *T*(23) = 8.05, *P*_*FWE*_ < 0.002. We also observed increased activation in area V4 in right inferior occipital gyrus with peak at MNI coordinate [30 −92 −2], *T*(23) = 10.96, *P*_*FWE*_ < 0.0001. There were no differences in BOLD amplitude between patients and controls at a standard family-wise error rate of *P*_*FWE*_ < 0.05.

### Brain mapping commonalities among patients and controls

We then used a conjunction analysis to identify regions active both during the perception of visual motion in general (social and non-social) as well as during social motion in particular (social minus non-social) in both patients and controls. This revealed two main clusters in the right hemisphere centred on motion-sensitive area V5 and the posterior superior temporal sulcus (pSTS) summarized in Table 3. Importantly, the activation of the pSTS during social motion conforms to the anatomical findings in the literature (Schurz et al., 2017). The time-series in these two regions, common to both patients and controls, were then used for the DCM analysis.

**Table 3.** Conjunction analysis: differences between social and non-social in regions that were also activated by motion in general

| *T* statistic | MNI coordinate | Anatomical region | Probabilistic atlas<sup>1</sup> |
| --- | --- | --- | --- |
| 16.12 | [46 -68 0] | Right middle temporal<br>Gyrus | hOc5 (V5) 52 %<br>hOc4la 47% |
| 18.06 | [-48 -70 0] | Left middle<br>occipital Gyrus | hOc5 (V5) 45 %<br>hOc4la 55 % |
| 5.61 | [54 -54 14] | Right superior<br>temporal sulcus | PGa (IPL) 45 %<br>PGp (IPL) 17% |

<sup>1</sup>Anatomical classification using the SPM anatomy toolbox (Eickhoff et al., 2005)

### Effective connectivity between V5 and pSTS in healthy controls

We then analysed the strength of extrinsic connections between V5 and pSTS in the right hemisphere, as well as changes in the inhibitory connectivity within each area. While the responses to motion in general were modelled as a driving input to V5, the responses to social versus non-social stimuli were modelled as a modulation (increase or decrease) of the connection strengths and hence constituted the experimental effects of interest. Bayesian model comparison of model evidences at the first level (Penny et al., 2010) revealed that the cortical network with changes in both extrinsic and intrinsic connections was the most frequent within healthy controls (Posterior model probability > 0.85 and protected exceedance probability > 0.99). This was confirmed by a Bayesian model comparison of the model evidences at the second level under parametric empirical Bayes (Posterior model probability > 0.99). Within this network, healthy controls had an increase in feedforward connectivity from V5 to pSTS (Posterior probability > 0.99) and a decrease in feedback connectivity (Posterior probability > 0.99). At the same time, there was a decrease in inhibitory influence within V5 (Posterior probability > 0.99) and an increase in inhibitory influence within pSTS (Posterior probability > 0.99).

### Effective connectivity in patients with schizophrenia compared to healthy controls

Using parametric empirical Bayes, we then analysed the strength of extrinsic and intrinsic connections in patients with schizophrenia compared to healthy controls. Again, Bayesian model comparison of model evidences at the first level (Penny et al., 2010) revealed that the full model was the most frequent across patients and controls (Posterior probability > 0.89 and protected exceedance probability > 0.999). This was confirmed by a Bayesian model comparison of the PEB model evidences at the second level (Posterior probability > 0.99). Within this network, patients had increased feedforward connectivity when they perceived social stimuli compared to their healthy controls (Posterior probability > 0.97).

### Patients with stronger positive symptoms have more disinhibition within pSTS

We then tested for the association between psychopathology and effective connectivity during social stimuli compared to non-social stimuli. Using parametric empirical Bayes, we compared the model evidence of a DCM with connection strengths explained by positive symptoms to the model evidence of a DCM explained by negative symptoms. Bayesian model comparison showed that between-patient differences in effective connectivity were better explained by positive symptoms than by negative symptoms. Inspection of the model revealed that patients who reported a higher degree of positive symptoms had a less influence of the inhibitory population onto the excitatory population within pSTS (Posterior probability > 0.99).

## Discussion

### High-functioning FES patients have aberrant brain connectivity

In this study, we used fMRI during the HCP social cognition paradigm (Barch et al., 2013) to test for differences in effective connectivity between patients with first-episode schizophrenia and matched controls. Our behavioral data summarized in Table 2. suggest that these first-episode patients were high-functioning in relation to previous findings where patients with schizophrenia typically performed 1-2 standard deviations below healthy controls on cognitive and social cognition tasks (Kern et al., 2004; Penn et al., 2008; Bora et al., 2009; Fioravanti et al., 2012; Savla et al., 2013; Fatouros-Bergman et al., 2014). This might be due to a short duration of illness, combined with a successful match of FES patients and controls. Despite their high level of functioning, our DCM results show aberrant feedforward connectivity in relation to matched controls. Crucially, we were able to show that between-patient variability in cortical inhibition within pSTS is associated with the severity of positive symptoms.

### Brain connectivity and predictive coding

Recent studies have shown differences in pSTS activation and functional connectivity with prefrontal cortex between patients with schizophrenia and healthy controls during different social perception paradigms (Backasch et al., 2013; Ciaramidaro et al., 2015; Mier et al., 2017; Jimenez et al., 2018; Okruszek et al., 2018). In line with our finding, Backash et al. showed an association between pSTS activation and delusional (positive) symptoms (Backasch et al., 2013). In contrast to these previous studies however, we show the association between psychopathology and brain functioning at the underlying neuronal level, as opposed to the level of observed BOLD responses. At the computational level, these differences in excitatory and inhibitory synaptic efficacy between patients and controls can be interpreted under predictive coding. In this framework, feedback connections encode prior beliefs in the form of predictions about hidden states in the world, such as the mental states of other agents that constitute theory of mind. By contrast, feedforward connections mediate the prediction errors that are inconsistent with these prior beliefs. The role of prediction errors is thus to update the representations of higher-level populations that provide Bayes-optimal predictions about the world. Our observation of stronger feedforward connectivity in patients fits well with current theories proposing that schizophrenia is associated with abnormally high levels of prediction error during perceptual inference (Kapur, 2003). Computationally, this increased level of feedforward influence may be compensatory in nature and reflect a state where prediction errors are weighted with an abnormally high level of precision.

### Aberrant encoding of precision in schizophrenia

In predictive coding, the precision with which a population of neurons represent hidden states in the world is thought to be encoded by their post-synaptic gain. The gain of a neuron represents its sensitivity to initiate a post-synaptic response following pre-synaptic input. The slope of this input-output relationship is controlled by the balance of excitation and inhibition and involves both fast AMPA receptor-mediated currents and voltage-dependent *N*-methyl-*D*-aspartate (NMDA) receptor mechanisms with slower time-constants (Isaacson and Scanziani, 2011). In schizophrenia, an imbalance in the precision with which prediction errors are encoded could arise from the gain control of glutamatergic NMDA receptor-mediated mechanisms by GABAergic interneurons (66). Evidence from non-invasive electrophysiology in humans has been reported by Schmidt et al. who observed an increase in feedforward connectivity within the auditory system (Schmidt et al., 2013) and Rosch et al. who observed a selective disinhibition within the superior temporal gyrus, both under NMDA-R blockade with ketamine (Rosch et al., 2018). Similar findings under pharmacological manipulation of NMDA-R function have been linked to psychosis (Adams et al., 2013; Friston et al., 2014; 2016). Our finding of disinhibition within pSTS in patients with positive symptoms concurs with these studies and adds to the growing evidence that psychosis is associated with an abnormal balance of excitation and inhibition (O’Donnell, 2011; Jardri and Denève, 2013).

### Future research

In this study, we used biophysical modelling of neuroimaging data to make inferences about the underlying mechanisms that mediate synaptic efficacy in the brain. This allowed us to identify pathophysiological differences in excitatory connectivity and local inhibitory mechanisms in patients with first-episode schizophrenia compared to healthy matched controls. We are currently planning more research integrating MRI with MEG and EEG in order to identify both differences and commonalities in pathophysiology across different subgroups of patients with schizophrenia, ranging from children at genetic risk of developing schizophrenia, first-episode patients and chronic schizophrenia.

### Replicability

Our reasons for using the HCP social cognition paradigm are twofold. First, it allowed us to replicate previous findings in the typical brain (Hillebrandt et al., 2014) using the exact same paradigm. Second, it allowed us to test for aberrant pathophysiology in a patient cohort using a standardized paradigm. Sharing of standardized stimulus paradigms and data analysis pipelines is thus absolutely essential for the replicability of neuroimaging findings in new datasets from both healthy and clinical cohorts across independent research sites.

## Data Availability

Anonimised MRI data are available upon request. Clinical and behavioural data are under GDPR protection.

## Acknowledgments

The project was funded with “Seed Money” from the Interacting Minds Center, Aarhus University. MJD is supported by VELUX FONDEN (00013930). We thank patients and psychiatric staff at the OPUS Clinic, Aarhus University Hospital, research assistants Maria Lotus Thai, Anna Sofie Lose Landert, Anne Katrine Kjærskov and Torben Lund at CFIN, Aarhus University.

## Author contributions

Conception of hypotheses: VFB, MJD, YZ and CDF. MRI data acquisition: VBF, LV and MJD. Neuropsychological testing: VFB and LV. Analysis of neuropsychological tests: VFB. Analysis of behavioural fMRI task: MJD. Analysis of fMRI data: MJD. MJD prepared figures. MJD and VFB wrote the manuscript with expert help from YZ and CDF.

